# Validity of markers and indexes of systemic inflammation in predicting mortality in COVID 19 infection : A hospital based cross sectional study

**DOI:** 10.1101/2021.03.30.21254635

**Authors:** B Archana, Shylaja Shyamsunder, Rinki Das

## Abstract

**Background:** COVID-19 is an ongoing global pandemic. It is a systemic infection with a significant impact on the hematopoietic and the immune system. In this study we aimed to evaluate the different inflammatory markers and indexes of systemic inflammatory response in predicting the mortality in patients with COVID 19.

**Methods:** In this cross sectional study, various inflammatory markers like D-dimer, CRP, serum ferritin, LDH and CBC derived indexes of inflammation were analyzed in predicting mortality in COVID 19 infection.

**Results:** We enrolled 302 COVID 19 patients who had a mean age of 54.51±15.39 yrs with 210 (69.5%) males. Among them 21% were asymptomatic and fever was the commonest among symptomatic patients. Majority of patients (66.7%) had no comorbidities and 20% had multiple comorbidities. On analyzing different hematological variables, survivors had statistically significant higher hemoglobin count, lymphocytes, monocytes, eosinophil and platelet count and lower leukocyte, neutrophil count. Inflammatory markers D-dimer, serum ferritin and LDH were significantly elevated among non survivors. Among the indexes of inflammation, only NLR showed significant higher values among non survivors.

All the inflammatory markers were able to predict mortality among the COVID 19 infected cases with a sensitivity and specificity of 85% and 65% for d dimer levels, 85% and 72% for serum ferritin, 85% and 72% for LDH, 85% and 51% for CRP levels respectively. Among the indexes of inflammation, validity of NLR was best in predicting mortality with 85% sensitivity and 51% specificity.

**Conclusion:** Abnormalities in peripheral blood parameters and increase in inflammatory markers are common findings in COVID 19 infection. NLR was best at predicting mortality followed by D-dimer and serum ferritin levels

**Contribution details:** 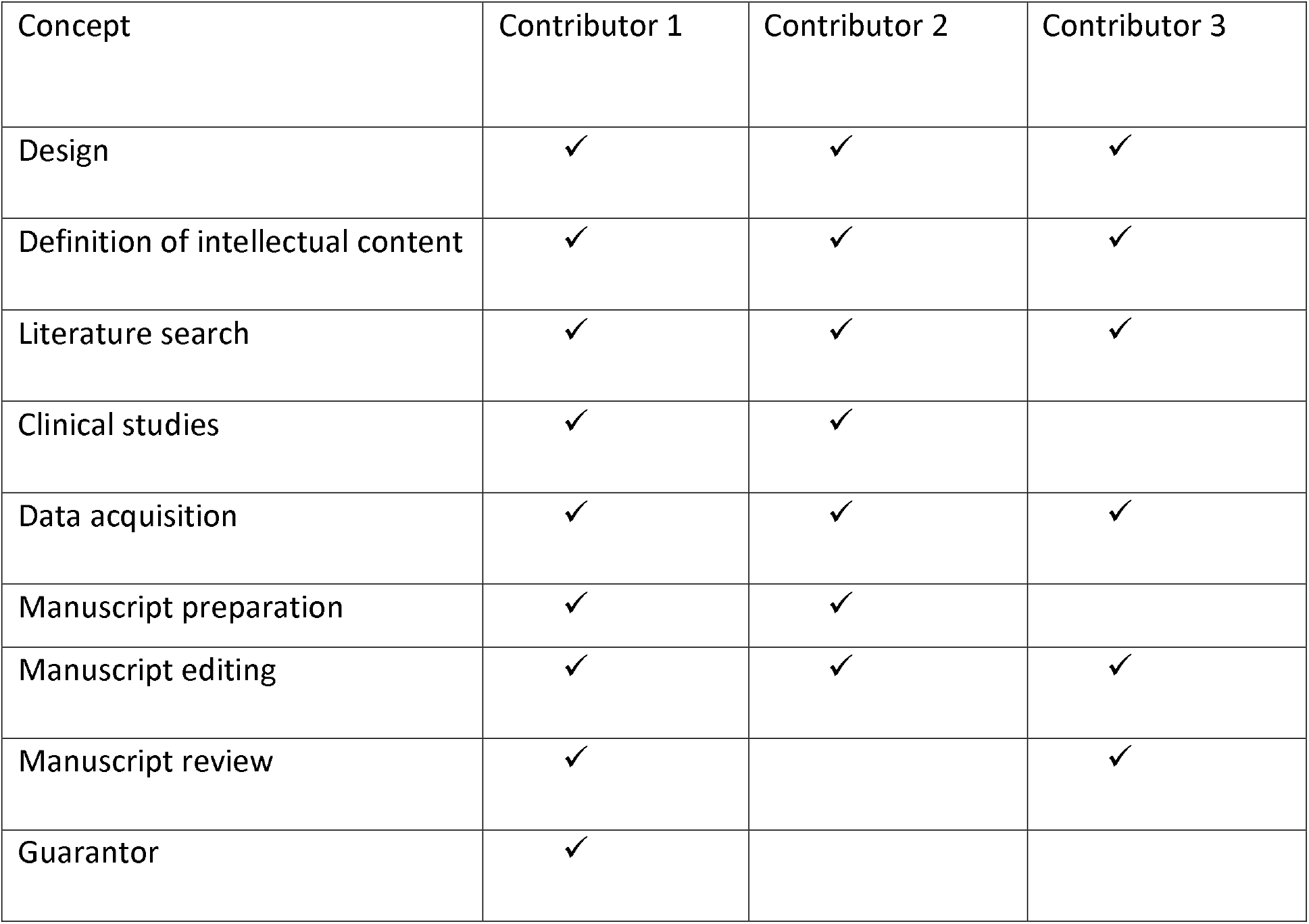

## Introduction

Coronavirus disease 2019 (COVID-19) caused by Severe acute respiratory syndrome coronavirus 2 (SARS-CoV-2) has rapidly spread over several months from an epidemic outbreak in Wuhan, China ^(1)^ into a pandemic, affecting patients across all age groups and geographic areas. Till date more than two million people have been succumbed to the infection. ^(2)^ The disease has a diverse course; patients may range from asymptomatic to those with respiratory failure, complicated by acute respiratory distress syndrome (ARDS). Every other organ has been affected by this virus including cardiovascular, respiratory, gastrointestinal, neurological, hematopoietic and immune system. ^(3–5)^ Nearly 20% of patients with COVID-19 become critically ill, with a high mortality ranging from 8.1% to 33%. ^(6,7)^ Older people and those with comorbidities are at increased risk of death from COVID-19.

Since COVID 19 is associated with high mortality rate and short time in the onset of ARDS after admission to hospital, ^(6)^ identification of early biomarkers of disease severity like CRP, D-dimer, LDH and serum ferritin might facilitate early aggressive treatment thereby reduces mortality and improves hospital resource allocation. Also the complete blood count (CBC) provides information regarding various cell types and morphological parameters, i.e., white blood count (WBC), lymphocytes, neutrophils, monocytes, platelet count and combined ratios of these parameters (indexes of inflammation) like neutrophil to lymphocyte ratio (NLR), platelet to lymphocyte ratio (PLR), monocyte to lymphocyte ratio (MLR), systemic inflammation response index (SII) and systemic inflammation response index (SIRI) can be used as marker for severity assessment of COVID-19 patients. ^(8, 9)^

The circulating biomarkers representing the immune system and inflammation have been considered as a prognostic indicator in COVID-19-positive patients and they can be easily performed and inexpensive. However their utility in predicting mortality is less explored.

## Material and Methods

This is a cross sectional study conducted at Bhagawan Mahaveer Jain hospital, Girinagar, Bangalore, from October 2020 to December 2020. All consenting consecutive patients with COVID 19 infection admitted to the hospital during the study period were enrolled in the study. Informed written consent was taken from the patients and the study protocol was approved by the Bhagawan Mahaveer Jain hospital Ethics Committee (BMJH/ECHR/29/2020). Patients were evaluated on arrival to the hospital and triaged to ward, high dependency unit or intensive care unit (ICU) as per clinical assessment. A focused history including age, gender, symptoms, duration of onset of symptoms, comorbidities were collected and base line saturation, general physical examination, systemic examination were done for all the enrolled patients and patients were categorized into mild (patients with mild symptoms without evidence of breathlessness and hypoxia), moderate [pneumonia with no signs of severe disease including SpO2 <94% (range 90-94%) on room air, respiratory rate>24/min] and severe category (with clinical signs of pneumonia plus one of the following: respiratory rate>30/min, severe respiratory distress, Spo2 < 90% on room air) accordingly.

Chest X ray and blood investigations including CBC, serum electrolytes, renal and liver function tests, inflammatory markers – CRP, D-dimer, serum ferritin, LDH were done at baseline. IL 6 and HRCT were done whenever indicated. Different CBC derived inflammatory indexes were calculated including neutrophil to lymphocyte ratio (NLR), platelet to lymphocyte ratio (PLR), monocyte to lymphocyte ratio (MLR), systemic inflammation response index (SIRI) (neutrophils X monocyte) / Lymphocytes and systemic inflammation index (SII) (neutrophils X platelet) / Lymphocyte.

## Statistical analysis

Data analysis was done using SPSS for Windows version 19.0 (IBM SPSS Statistics). Study variables were presented using descriptive measures like mean ± SD or median with minimum-maximum range. Continuous variables were compared by Mann Whitney test and frequency proportions by the Chi-square test or Fisher’s exact test. Statistical significance was set at P < 0.05. Receiver operating characteristics (ROC) curve analysis was performed to estimate optimal cut off values, sensitivity and specificity for prediction of the outcome of interest

## Results

We enrolled 302 patients in the study. Mean age of the study patients was 54.51±15.39 yrs with 210 (69.5%) males. Mean duration of symptoms was 6.03 days. A significant number of patients were asymptomatic (21%) and among the symptomatic fever (84%) was the most common presenting complaint, followed by cough (59%), loss of smell and taste (40%), breathlessness (30%), body ache and fatigue (17%) and sore throat (6%). Majority of patients (66.7%) had no comorbidities and 20% had multiple comorbidities. Diabetes mellitus (26.6%) was the commonest co morbidity followed by hypertension (21.3%), ischemic heart disease (6%), COPD (2%) and asthma, OSA, malignancy, seizure disorder, CVA 0.6% each. Out of 302 patients, 236 (78.1%) patients had mild, 44 (14.5%) had moderate and 22 (7.2%) had severe infection (p=<0.001). Out of 302, 287 (95%) were successfully treated and discharged home and 15 (4.9%) succumbed to the infection.

Statistically significant difference was seen between the survivors and non survivors across different hematological variables (Table 1). Mean age of survivors was lesser compared to non survivors (p=0.021). Similarly baseline mean Spo2 was higher (p<0.001), mean hemoglobin count was higher (p=0.023) among the survivors. Also survivors had lower total count (p=0.01), lower neutrophil count (p=0.009), higher lymphocyte count (p=0.044), higher monocyte count (p=0.04), high eosinophil count (p=0.12), high platelet count (p=0.36). There was no significant difference in electrolytes, AST, ALT between the two groups. However significant difference was seen in creatinine levels (p=0.03) between them.

**Table 1:** Demographic, clinical and hematological features of COVID-19 survivors and non-survivors.

| Variables | Survivors (n= 287) | Non survivors (n=15) | p value |
| --- | --- | --- | --- |
| Age (years) | 53.87±15.42 | 67.57±6.90 | 0.02 |
| Gender (male %) | 200 (69.6) | 10 (66.6) | 0.48 |
| Duration of symptoms (days) | 5 (16) | 5 (19) | 0.89 |
| Baseline saturation (%) | 94.66±3.90 | 84.29±14.06 | <0.001 |
| Co morbidities (n %) | 29 (20.2) | 5 (71.4) | 0.03 |
| Hemoglobin (g/dl) | 13.20±1.86 | 11.51±2.52 | 0.02 |
| WBC (X 10<sup>3</sup>/cumm) | 7.21 (2.46-20) | 10.96 (6.4-15.55) | 0.001 |
| Neutrophils (%) | 69 (39-98) | 90 (68-93) | 0.009 |
| Lymphocytes (%) | 23 (1-50) | 8.2 (4.3-30) | 0.04 |
| Monocytes (%) | 4.3 (0.1-39) | 1.4 (0.4-4.6) | .004 |
| Eosinophil (%) | 0.7 (0.0-20) | 0.0 (0.0-2) | 0.01 |
| Platelets (X 10<sup>5</sup>/cumm) | 2.4 (0.95-4.95) | 2 (0.97-2.8) | 0.03 |
| AST (u/l) | 29 (10-125) | 40 (20-50) | 0.38 |
| ALT (u/l) | 29 (10-140) | 44 (13-52) | 0.58 |
| Serum creatinine (mg/ml) | 0.7 (0.1-2) | 2 (1-4) | 0.00 |
| CRP (mg/L) | 18 (0.2-329) | 62 (5-314) | 0.08 |
| D-dimer (ng/ml) | 307 (32-1000) | 604 (395-1803) | 0.02 |
| Serum ferritin (ng/ml) | 222 (5.54-3000) | 480 (310-2000) | 0.02 |
| LDH (u/l) | 239 (48.8-938) | 320 (218-964) | 0.03 |

WBC- white blood cells, AST- aspartate aminotransferase, ALT- alanine transaminase, CRP- C reactive protein, LDH- lactate dehydrogenase. All continuous variables are reported as mean ± standard deviation, medians and minimum – maximum range. Statistical significance set at 0.05.

On analyzing the inflammatory markers, statistically significant difference was found between two groups in D-dimer levels (307 vs 604, p=0.021), serum ferritin (222 vs 480, p=0.023) and serum LDH levels (239 vs 320, p=0.036). Even though CRP levels were much lower in survivor group (18 vs 62, p=0.08), it was not statistically significant.

Combined blood cell count indexes of inflammation were analyzed between two groups (Table 2). Non survivors had higher values of NLR (8.40 vs 2.95, p=0.04), PLR (208 vs 133, p=0.2), MLR (20.54 vs 14.1, p=0.16), SIRI (1607.7 vs 928.5, p=0.48), SII (11.53 vs 9.64, p=0.52). However statistically significant difference was seen only for NLR.

**Table 2:** Indexes of systemic inflammation in COVID-19 survivors and non-survivors.

| Indexes | Survivors (n= 287) | Non survivors (n=15) | p value |
| --- | --- | --- | --- |
| NLR | 2.95 (0.3-98) | 8.4 (2.8-21) | 0.04 |
| PLR | 133 (74-407) | 208 (92-4164) | 0.2 |
| MLR | 14.1 (8.6-64.8) | 20.5 (12.4-2006) | 0.16 |
| SIRI | 928.5 (543-22534.3) | 1607.7 (736.4-3457.6) | 0.48 |
| SII | 9.6 (4.6-230.5) | 11.5 (6.2-24.6) | 0.52 |

NLR - neutrophil to lymphocyte ratio, PLR - platelet to lymphocyte ratio, MLR - monocyte to lymphocyte ratio, SIRI - systemic inflammation response index (neutrophil\* monocyte to lymphocyte ratio), SII - systemic immune-inflammation index (neutrophil\*platelet to lymphocyte ratio). All variables are reported as median and minimum - maximum range. Statistical significance set at 0.05.

A receiver operating characteristic curve (ROC) analysis was carried out to find out cut off values with highest sensitivity, specificity and validity of markers and indexes of inflammation in predicting mortality. As seen from ROC figure (Figure 1), validity of NLR was best in predicting mortality among the COVID 19 infected. The AUC (.787) was highest for NLR compared to other indexes (Table 3). 85% sensitivity and 51% specificity was found for NLR at cut off value of 2.98. Similarly 42% sensitivity and 49% specificity was found for PLR at 205, 28% sensitivity and 49% specificity for MLR at 17.2, 57% sensitivity and 71% specificity for SIRI at 1523 and 71% sensitivity and 53% specificity for SII at 10.8.

**Table 3.** Receiver operating characteristics (ROC) curves, sensitivity and specificity of markers and Indexes of systemic inflammation in predicting mortality in COVID-19 infection

| Parameters | AUC | Cut off | Sensitivity (%) | Specificity (%) |
| --- | --- | --- | --- | --- |
| D-dimer | .759 | >532 (ng/ml) | 85 | 65 |
| Serum ferritin | .755 | >312 (ng/ml) | 85 | 72 |
| LDH | .735 | >279 (u/l) | 85 | 72 |
| CRP | .696 | >18.4 (mg/L) | 85 | 51 |
| NLR | .787 | >2.98 | 85 | 51 |
| PLR | .358 | >205 | 42 | 49 |
| MLR | .346 | >17.2 | 28 | 49 |
| SIRI | .578 | >1520 | 71 | 71 |
| SII | .571 | >10.8 | 71 | 53 |

CRP- C reactive protein, LDH- lactate dehydrogenase, MLR- monocyte to lymphocyte ratio, NLR - neutrophil to lymphocyte ratio, PLR- platelet to lymphocyte ratio; SII - systemic immune-inflammation index, SIRI - systemic inflammation response index

**Figure 1:**
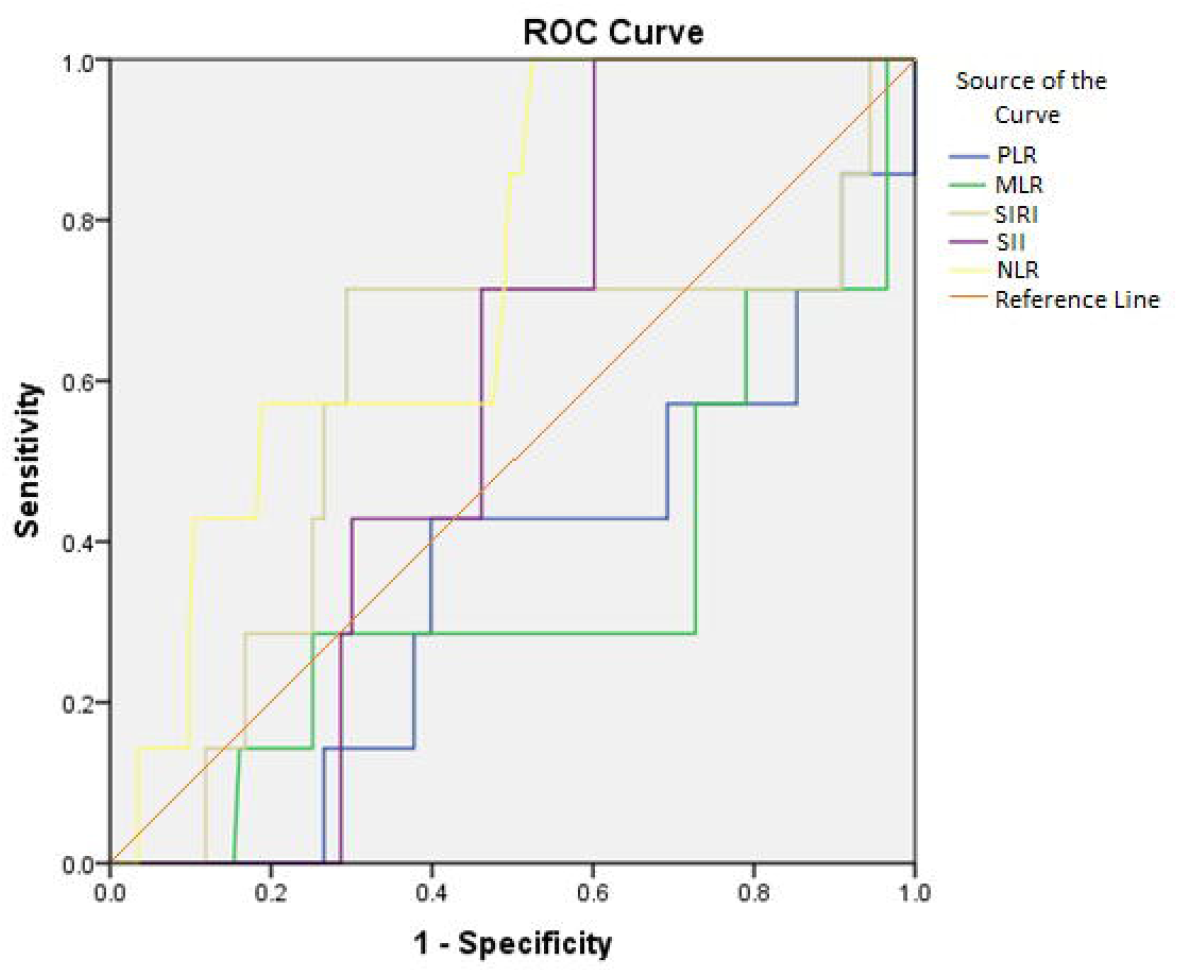
ROC curve for indexes of systemic inflammation

All the inflammatory markers were able to predict mortality among the COVID 19 infected cases (Figure 2). However AUC (.759) was highest for d dimer compared to other markers (Table 3). 85% sensitivity and 65% specificity was found for D-dimer at cut off value of 532. Similarly 85% sensitivity and 72% specificity was found for serum ferritin levels at 312, 85% sensitivity and 72% specificity for LDH at 279, 85% sensitivity and 51% specificity for CRP at 18.4.

**Figure 2:**
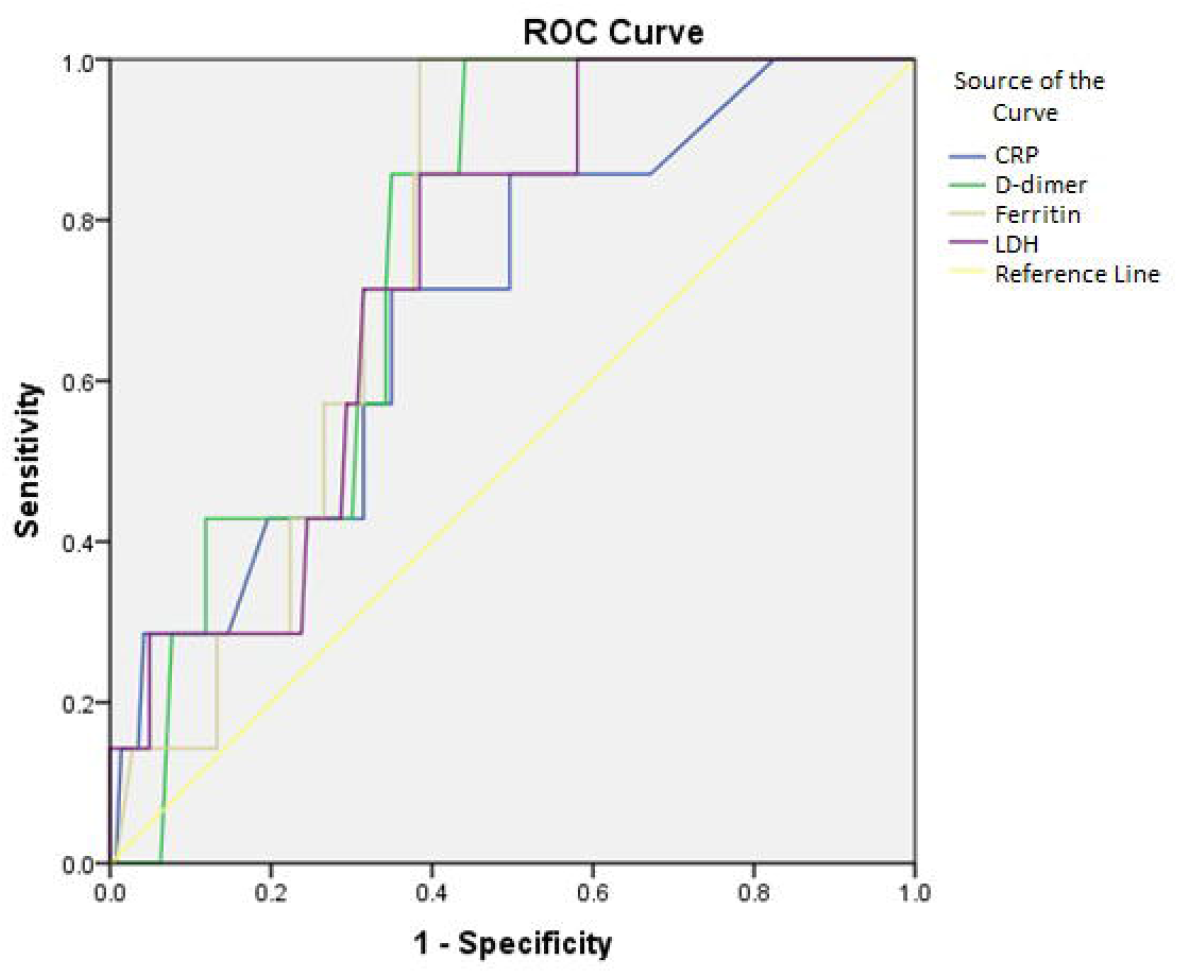
ROC curve for inflammatory markers

## Discussion

Since the outbreak of 2019-nCoV pneumonia in December 2019, >110 million people are infected worldwide with >10 million infections in India and the number continues to grow with case fatality rate of 2.19% and 1.44% worldwide and in India respectively. ^(2)^ The current difficulty is the lack of medical resources, especially critical care resources. If cases with higher risk of mortality can be identified as early as possible, risk stratification can help in better utilization of insufficient medical resources and might reduce mortality. There is insufficient data in the literature on predictors of mortality in COVID 19 infection at the time of admission to the hospital. Our study provide several important insights about the sensitivity and specificity of various baseline makers and indexes of inflammation in predicting mortality in these patients.

Clinical and demographic characteristics of 302 patients enrolled in our study was similar to those described in recent literature. ^(10)^ Fever was the commonest symptom followed by cough, loss of taste & smell and breathlessness which was similar to other studies. ^(6,10)^ As expected, non survivors were of higher age group compared to survivors along with lower baseline saturation and more co morbidities. ^(11–13)^ Diabetes mellitus was the commonest co morbidity followed by hypertension and ischemic heart disease. We did not find any difference in the duration of symptoms between the two groups.

CBC can be easily performed and is inexpensive. Parameters such as leukocytes, neutrophils, lymphocytes, monocytes, eosinophil and platelet count, individually and in combination can be used as indexes of systemic immune response. In our study, non survivors had higher leukocytes, neutrophils and lower lymphocytes, monocytes, eosinophil and platelet count which was similar to findings of other studies. ^(10, 14, 15)^

Circulating leukocytes respond to stress by increasing neutrophils and reducing lymphocytes. Lymphopenia is extensively studied as a good predictor of severe disease and worse outcomes in COVID 19 pneumonia. Causes of lymphopenia in COVID 19 pneumonia are several like (1) SARS-CoV-2 mainly infects T lymphocytes which expresses the ACE2 receptor on their surface and leading to their lysis. ^(16)^ (2) Lymphocyte apoptosis caused by increased levels of cytokines in response to systemic inflammation. (3) Atrophy of lymphoid organs, including the spleen which impairs lymphocyte turnover. ^(17)^ (4) Coexisting lactic acid acidosis in some at risk patients may also inhibit lymphocyte proliferation. ^(18)^ A high leukocyte count is common in critically ill patients because damaged cells induce innate inflammation in the lungs, which is largely mediated by proinflammatory macrophages and granulocytes and neutrophilia may also indicate superimposed bacterial infection, a finding described by Fan et al in ICU hospitalized COVID 19 patients. ^[19]^ Thrombocytopenia which was more prominent among non survivors in our study may be due to abnormal coagulation function leading to increased platelet consumption and decreased platelet number. In addition, increased damage to the ACE2-receptor-rich kidney tissue ^(20)^ in SARS-CoV-2infection by various inflammatory factors, can lead to reduced erythrogenesis and increased destruction of RBC, leading to anemia, which was evident among non survivors in our study.

NLR is a widely used marker for the assessment of the severity of bacterial infections and the prognosis of patients with pneumonia and tumor. Various studies have shown that NLR positively correlates with the risk of COVID-19 and also elevated NLR is an independent prognostic biomarker that affects progression of pneumonia in COVID-19 patients. ^(15, 21, 22)^ In our study, we tried to analyze various peripheral blood combined parameters in predicting the mortality in COVID 19 pneumonia. Even though all the combined parameters were elevated among non survivors, only elevation in NLR was statistically significant compared to others like MLR, PLR, SIRI and SII, which is contrary to the findings of Qu R et al ^(23)^ and Fois A G et al ^(24)^ where PLR and SII predicted mortality in COVID 19 patients. The area under ROC curve was largest for NLR at values > 2.98 with higher sensitivity and specificity in predicting mortality. This shows that along with immune system dysfunction which is evident in COVID 19 infection, superadded bacterial infection may also play an important role in COVID 19 outcome.

As knowledge of COVID-19 expands, various inflammatory markers which are measurable by readily available and inexpensive tests, have been suggested for assessing severity of disease. Our study showed that elevated CRP, ferritin, D-dimer and LDH levels were significantly associated with in hospital death in COVID 19 pneumonia.

Coagulation disorders are relatively frequently encountered among COVID-19 patients, especially among those with severe disease. Various studies have highlighted the role of D dimer in assessing the severity and mortality in COVID 19 pneumonia. ^(11, 25)^ Elevated D-dimer and fibrinogen degradation products in non survivors have been linked to coagulation activation, dysregulated thrombin generation, impaired natural anticoagulants, and fibrinolysis. ^(26)^ Also, several critically ill patients have been reported to develop coagulopathy, antiphospholipid antibodies, and increased arterial and venous thrombotic events such as cerebral infarction. ^(27)^

CRP, an acute phase reactant that is increased in a wide range of inflammation and infection has been found to be increased in 75%-93% of patients with COVID-19 infection, particularly in severe disease. ^(28)^ These above findings were also evident in our study. We also found COVID 19 non survivors had higher levels of serum ferritin which is a surrogate marker of stored iron and increase in inflammation, similar to findings by Zeng F et al. ^(29)^ Manson JJ et al in their study reported that COV HI criteria (CRP >150 mg/L or ferritin levels >1500 μg/L) at admission was associated with increased mortality. ^(30)^ LDH was increased in non survivors in our study which was in agreement with other studies which showed elevated LDH was common in COVID-19 patients in the ICU setting and indicated a poor outcome. ^(19, 28)^ The results of our study also showed that all these parameters had high predictive values in terms of mortality in COVID 19 pneumonia. The area under ROC curve of D-dimer was the largest with high sensitivity and specificity, followed by the AUC of serum ferritin, LDH and CRP. All of these highlight that an over exuberant inflammatory response is associated with mortality in COVID-19.

Our study has few limitations. We studied only the baseline parameters in predicting mortality rather than a dynamic monitoring. These results needs to be validated in a large cohort.

## Conclusion

COVID 19 is a systemic infection with a significant impact on the hematopoietic and the immune system. Abnormalities in peripheral blood parameters and increase in inflammatory markers are common findings in COVID 19 infection. NLR was best at predicting mortality followed by D-dimer and serum ferritin levels. Careful evaluation of these parameters at baseline can assist the clinicians in early risk assessment and further close monitoring may probably reduce the mortality.

## Supporting information

Ethical approval letter

## Data Availability

All the data is available

## Acknowledgment

Authors would like to thank Dr Siddalingappa Hugara for helping in statistical analysis.

