## Supplementary material for "Validity of markers and indexes of systemic inflammation in predicting mortality in COVID 19 infection : A hospital based cross sectional study": Ethical approval letter

ETHICS COMMITTEE ON HUMAN RESEARCH
BHAGWAN MAHAVEER JAIN HOSPITAL
A UNIT OF BHAGWAN MAHAVEER MEMORIAL JAIN TRUST
Millers Road, Vasanthnagar, Bangalore - 560 052.


**BMJH/ECHR/29/2020** **August 6, 2020**

To,
**Dr. Archana B**
Department of Pulmonary Medicine
Bhagwan Mahaveer Jain Hospital,
Girinagar, Bangalore

Dear Dr. Archana B

**Sub : Ethics Committee Approval – Reg**

The BMJH Ethics Committee on Human Research, Millers Road, Vasanthnagar, Bangalore 560 052, has reviewed and discussed the said Cross Sectional Study title ***Validityof markers and indexes of systemic inflammation in predicting mortality in COVID 19 infection – A cross sectional study"*** conducted at Bhagwan Mahaveer Jain Hospital, Girinagar Branch, Bangalore, and submitted by you to The BMJH Ethics Committee for approval for publication in the Indian Journal Of Critical Care Medicine.

The BMJH Ethics Committee has reviewed the same and Approval has been accorded to you to submit the Study Paper to the Indian Society Of Critical care Medicine for publication in the Indian Journal Of Critical Care Medicine.

It is understood that this is not a clinical trial and there is no financial implication in the conduct of this study.

You are requested to kindly maintain confidentiality of patient identifications while submitting your paper for publication

I hereby confirm that this Ethics Committee is organized and operates according to ICH- GCP, Schedule "Y" Guidelines and the applicable laws and regulations.

Yours sincerely,

**DR. (WG CDR) M. D. MARKER** MBBS, MHA
**MEMBER SECRETARY,**
Bhagwan Mahaveer Jain Hospital Ethics Committee,
Bhagwan Mahaveer Jain Hospital
Millers Road, Vasanthnagar
Bangalore – 560 052

Member Secretery
Ethics Committee on Human Research
Bhagwan Mahaveer Jain Hospital
Miller's Road, Vasanthnagar
Bangalore-560 052
